# “Implementing a policy is something else”: Governance of complex health information systems in Tanzania

**DOI:** 10.1101/2024.08.15.24312044

**Authors:** Regine Unkels, Elibariki Mkumbo, Ntuli A Kapologwe, Fatuma Manzi, Claudia Hanson, Helle Mølsted Alvesson, Andrea B Pembe

**Affiliations:** Department of Global Public Health, Karolinska Institutet, Stockholm, Sweden; System, Policy and Economic Evaluations, Ifakara Health Institute, Dar es Salaam, Tanzania; Health, Social Welfare & Nutrition Services, President’s Office - Regional Administration and Local Government, Dodoma, Tanzania; Department of Disease Control, Faculty of Infectious and Tropical Diseases, London School of Hygiene and Tropical Medicine, London, United Kingdom; Department of Obstetrics and Gynaecology, Muhimbili University of Health and Allied Sciences, Dar es Salaam, Tanzania

**Keywords:** Health Management Information Systems, Health Policy, Complex Adaptive Systems, Tanzania

## Abstract

**Introduction:** Health management information systems are crucial for a country’s health service planning and monitoring. Research indicates that generated data is often of low quality or not used for decision-making in low-resource settings. Digitalization potentially alleviates these problems, but scale-up in these countries is hampered by unreliable availability of resources.

We aimed to understand how health policymakers perceive and experience working with data, data systems and the introduction of digital technology related to the governance of health management information systems in Tanzania.

**Methods:** We conducted 16 interviews with national, regional and district health care managers experienced in using health information systems in Tanzania. Reflexive thematic analysis was used. Themes were developed underpinned by complexity theory and M. Lipsky’s theory of street-level bureaucracy.

**Results:** Health care managers experienced challenges in health management information system governance in an unpredictable environment. Different power practices for system governance and implementation were used: Institutional power was applied to areas with existing international guidance and strategic examples. Subnational managers contextualized implementation through discretionary power practices where uncertainties prevailed. This led to transformed agendas in some cases, but also allowed for innovations to make policies work.

**Conclusions:** Acknowledging the complexity of health management information system governance with constant adaptation can allow policymakers and senior managers to direct discretionary power where policy implementation would otherwise fail in the Tanzanian context. This can be achieved by identifying a set of social values around data processes that resonates with all actor groups and may support governance of this complex system.

## 1. INTRODUCTION

Governance of health management information systems (HMIS) is crucial to ensure that good quality routine health data can be readily used for improving and maintaining health system performance (1, 2). Over the past 15 years, however, challenges with data quality and use, especially in low- and middle-income countries were documented (3-9). The imperative to improve this situation has fostered the digitalization of HMIS through the *District Health information System-2* (DHIS-2) in many low-and-middle-income countries (10). Evidence suggests that digitalized HMIS can improve data use (11) and alleviate calculation and transfer errors through automatization (11). In settings like Tanzania however, primary data is currently collected by hand in large paper registers and later digitized manually into DHIS-2 by a different person. These processes are complex (12) and may lead to erroneous data at several points in the process (9, 13).

Research from Tanzania and similar settings, on the other hand, indicates that health care providers (HCP) and their immediate supervisors may also manipulate data to maintain social relationships in a working environment that challenges good performance (12, 14-16). This has implications for data quality that could be amended by strong governance.

Tanzania has made great strides to create HMIS structures at policy level in the last 15 years. A national digital health strategy is in place (17) and the country is one of few that developed a road map for the digitalization of the health sector together with an enterprise architecture for digital governance in this area (18, 19). Increasingly, research on policymaking and policy implementations reveals the complexity of health governance systems and indicates that policy implementation is not only shaped by top-down guidance but also by a variety of actors at different implementation levels (20, 21).

The aim of this study was therefore to understand how health care policymakers at different health system levels perceived and experienced governance of health management information systems and the introduction of digital technology to support data quality and use.

## 2. THEORETICAL APPROACH

We applied complexity theory acknowledging the complex and interdependent nature of health policymaking and governance and of health management information systems (12, 21-25). *Complexity theory* describes biological and human networks as perpetually self-organizing systems in reaction to evolving external and internal environments and through adaptive learning processes within the systems (26). The theory defines modern health care systems as multi-component, non-linear, unpredictable, and adaptive networks of multiple interdependent agents (25). *Social constructivism* underlines the transactional nature between different actor groups within health policy and implementation and the importance of actor relationships rather than of individual actors (27). Barassa et al depicted lower-level hospitals in Kenya as complex adaptive systems. The authors described how priority setting and resource allocation were influenced by resource scarcity and low managerial capacity where revenue maximization emerged as an adoptive though unintended measure to keep the hospitals running (28). These findings relate to Michael Lipsky’s *theory of street-level bureaucrats* which proposes that frontline actors in a bureaucratic system may use discretion in interpreting policies, and their positional power to implement policies in everyday work (29). The theory has been applied to other qualitative health policy evaluations in sub-Saharan countries including Tanzania (20, 30, 31). These studies described how lower-level health care providers exercise discretionary power practices in the interpretation of health policy. Those practices often resulted in unintended or even negative outcomes for policy implementation e.g. on using their own rules for decision-making, not following standard guidelines or through inaction.

While complexity theory and social constructivism informed study design and analysis, we applied M. Lipsky’s *theory of street-level bureaucrats* to the refining of themes and the overarching theme during later stages of thematic analysis to include policy and power dimensions as well as the social and system perspective (24, 32).

## 3. METHODS

We report results from a qualitative study based on social constructivism using 16 in-depth-interviews with Tanzanian health care managers from national and subnational level and involved in governance of routine health data. We applied the *Consolidated Criteria for Reporting Qualitative Studies* (COREQ) (33) and the *Recommendations on Quality Practice and Reporting on Thematic Analysis* by Braun and Clarke (34).

### 3.1. Setting

Tanzania’s health system provides primary health care in dispensaries, health centers and district hospitals under the *President’s Office - Regional Administration and Local Government* (PO-RALG). Advanced care is delivered at regional and zonal referral hospitals under *Ministry of Health* (MoH) and tertiary specialized referral hospitals in bigger municipalities.

Facility-based routine health data is collected by HCPs in printed registers and is subsequently digitized manually in DHIS-2. Approximately 120 partly independent data systems operated in Tanzania in 2016, mostly created for data collection for vertical programmes (35). The Tanzanian Government has progressively put legislation and guidance in place to govern health sector digitalization (17, 18, 36). Currently three main public health data systems exist including DHIS-2 (12). These systems are partly integrated but have limited interfaces with other administrative data systems or vertical disease-specific systems.

At national level both PO-RALG and MoH entertain a dedicated unit for *Information Communication Technology* (ICT) including HIS governance. At subnational level managers in *Council Health Management Teams* (CHMTs) supervised by *Regional Health Management Teams* (RHMTs) oversee health care and HIS implementation.

This research was part of a larger study (*Action Leveraging Evidence to Reduce Perinatal Mortality and Morbidity in sub-Saharan Africa, ALERT*) to develop and evaluate an intervention to improve intrapartum care in four hospitals in Southern Tanzania through a health system strengthening lens. The core components of the ALERT intervention are data-driven health service quality improvement, governance and accountability mechanisms. (37).

### 3.2. Sampling and Recruitment

Purposive sampling was used for maximum variation, informed by the concept of *information power* (38). We selected eligible managers based on a set of inclusion criteria (Table 1) and recommendations by organizations working on health care digitalization, to facilitate the inclusion of varied views, in-depth knowledge and experience among managers. Potential managers were contacted by phone and email; written and oral information about the study and answers to arising questions were provided. After agreement to participate, appointments according to managers’ conveniences were arranged. All 17 contacted managers agreed to be interviewed and 16 of these eventually participated. One senior national manager cancelled several appointments due to a series of equally important meetings he had to participate in.

**Table 1.** Inclusion Criteria.

| Inclusion criteria | Exclusion criteria |
| --- | --- |
| Senior or mid-level managers/policy makers in the health sector of Tanzania | Managers or policy makers working in other public sectors than health care |
| Working at MoH or PO-RALG national level, or regional/district level | Not currently working for MoH or PO-RALG at national, regional or district level |
| Previous or current experience with using data from DHIS2 or other digital systems collecting routine health information<br>OR<br>previous or current experience in piloting, introduction or scale-up of new digital systems to collect routine health information | No current or previous involvement in either piloting, introduction, scale-up or data use of digital systems collecting routine health information |

*PO-RALG= President's Office - Regional Administration and Local Government; MoH=Ministry of Health; DHIS-2=District Health Information System-2.*

### 3.3. Data Collection

Topic guides were based on previous research on HMIS data collection in Tanzania (12, 23) and grounded in complexity theory and social constructivism (27, 39). The included topics were i) previous experience with HMIS, ii) perceptions on accountability, data quality and use, iii) personal involvement in design, supervision and application of HMIS and digital systems of data collection, among others. They were developed in English, translated into Kiswahili and reviewed for emerging topics after each interview.

Data collection took place in October 2022 in four district and regional capitals of Southern Tanzania and in January 2023 in Tanzania’s capital Dodoma. RU conducted all interviews, lasting 45 to 60 minutes on average. Two managers refused recording, but allowed notetaking, all others were recorded after consent. Most interviews were conducted in managers’ offices in Kiswahili or English as per participant’s choice. Audios were transcribed verbatim in Kiswahili or English by EM. Relevant quotes from Kiswahili transcripts were translated into English.

We interviewed eight managers from subnational (CHMT and RHMT) and eight managers from national level (PO-RALG and MoH) (Table 2). Only two managers were female, and 14 were male. This reflects the gender distribution related to ICT and HMIS in Tanzania. Participants’ ages ranged from age group 20-30 to over 50 years.

**Table 2.** Demographic Characteristics of Managers.

| Characteristics | Category | Managers (n= 16) |
| --- | --- | --- |
| Age | 20-30 | 1 |
|  | 31-40 | 6 |
|  | 41-50 | 5 |
|  | ≥51 | 4 |
| Gender | Female | 2 |
|  | Male | 14 |

### 3.4. Data analysis

We conducted analysis along the six phases of thematic analysis described by Braun and Clarke (40): RU and EM discussed impressions from data collection and transcription and read transcripts several times, then coded four transcripts independently to include variations in researchers’ positionality. RU then coded the entire data set inductively, while writing memos. During several rounds of coding, clusters of shared meaning were identified, discussed during regular peer checks with HMA and EM, then potential themes were developed further iteratively moving between i) data, codes and themes and ii) reflective sessions with research team members. Complexity in health care and *street-level bureaucracy* were used to inform the construction of themes.

### 3.5. Reflexivity

The research team included early career researchers (RU) and experienced researchers (EM, FM, ABP, NAK, CH and HMA) from middle- and high-income country institutions. RU, NAK, ABP and CH have medical backgrounds. HMA is a medical anthropologist, EM a social scientist and FM an economist with experience in stakeholder management. Three co-authors are male and four as female. Three team members are of European descent (RU, CH, HMA), all other members are from Tanzania.

RU, the first author, speaks fluent Kiswahili and has work experience in the study area. All research team members have contextual knowledge about the Tanzanian health system, HIS and stakeholder management. All members have access to the data.

### 3.6. Ethical considerations

Clearance was obtained in Tanzania from the Institutional Ethics Review Board of *Muhimbili University of Health and Allied Sciences* (MUHAS-REC-02-2022-975), from the *National Institute for Medical Research* (NIMR/HQ/R.8a/Vol IX/4009 and NIMR/HQ/R.8c/Vol I/2483) and from the *Swedish Ethical Review Authority* (2020-01587). Written informed consent from managers was obtained.

## 4. RESULTS

We report results along the three themes identified during analysis: i) “Shaping HMIS design through agenda-setting”, ii) “Shaping HMIS design through decisions”, iii) “Shaping HMIS design through connection” and nine sub-themes, all linking to one overarching theme called “Shaping HMIS governance using stick and carrot” (Figure 1). Quotes are used to illustrate findings where appropriate.

**Figure 1.**
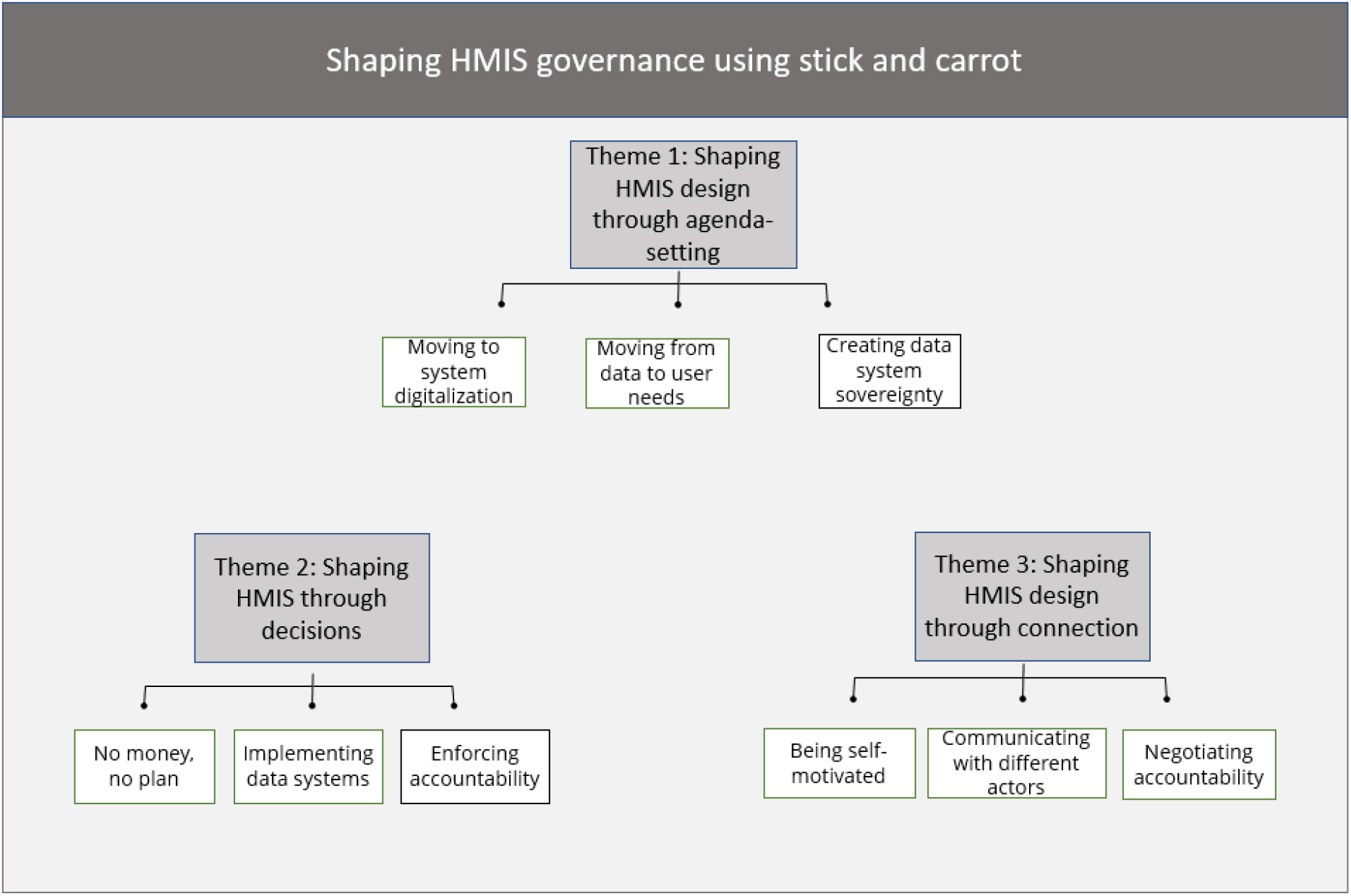
Themes and Sub-Themes (HMIS= Health management information system)

National and subnational managers (region and district) had different tasks within the HMIS, still they shared a lot of experiences and tacit knowledge as actors within the same organizational culture. Where we found important differences in power expressions or sensemaking, we are referring to the respective sub-group of the managers.

### 4.1. Theme 1: Shaping HMIS design through agenda-setting

#### 4.1.1. Moving to system digitalization

Digitalization of health data was reportedly high on the national agenda and was equally visible in elaborations of subnational managers. Managers described how digitalization, especially through electronic medical records, could improve data quality and indirectly accountability of primary data collectors.

> *“The day we remove manual data entry for every facility we will have good quality data. This is an important aim of the ministry [MoH] and in the health sector strategic plan 5 and we [therefore] need to digitalize the whole lower level of the health sector*.*”* (IDI 9 national level)

The quote depicts the high expectations of national managers in terms of data quality through digitalization. This extract also provides insights into underlying presumptions of the participant, about the power of digitalization to also address behavioral factors in data processing. Managers also explained that currently HCPs had little computer knowledge, especially in lower-level facilities. They hoped that time would solve this through the employment of younger HCPs with improved digital skills, assuming they would better understand benefits from digitized data.

#### 4.1.2. Moving from data to user needs

Data quality was also important from the national perspective because high-quality data was seen as the basis for effective data use nationally but also internationally. Managers’ emphasis on data quality was based on their familiarity with the clear guidance in the international data quality assessment (DQA) tool from World Health Organization (WHO) which was also implemented in Tanzania.

> *“If you look at strategic planning, the objective is to improve availability of good quality data so that it can assist us to evaluate ourselves, to improve, to enable us to see how we can measure ourselves as a country*.*”* (IDI 9 national level)

In contrast, there were no internationally or nationally agreed measures to improve data use. Especially subnational managers thought that use would improve data quality, through HCPs experiencing the impact of low-quality data on available resources at facility level.

> *“It should have been quality first then use, but now we think, they should start using it [data] first. Once they start using it [data], they will say “no, this [data] is not correct*.” (IDI 15 national level)

National managers described politicians as their most important end users of data. The importance of this user group may have also determined the focus on performance monitoring and planning, which could also explain the emphasis on numeric data and on quality over data use.

> *“Nowadays they understand the importance of data, because in the past five or six years, we had leaders where you could not do anything without data. So, you write a report, they [leaders] will tell you: “[where is the] data”? You want something, they will tell you “Convince me with numbers” … So, this has even built a national level culture of data use*.*”* (IDI 15 national level)

#### 4.1.3. Creating data system sovereignty

Managers mentioned that system roll out, data quality assurance, but especially digitalization were often donor dependent. National-level managers emphasized efforts to create an HMIS that was entirely owned by the Government of Tanzania by i) owning the agenda, ii) driving system integration and iii) applying institutional power for appropriation of newly developed tools. Subnational managers’ vision of data sovereignty seemed to be rather about their agency getting things done with the support of donor organizations. This may indicate a loss in translation of the national agenda at subnational level.

> *“So sometimes you can get a place [in the car of another supervision team] based on funds availability for one round and where we have donors, we do it [DQA] twice. So yes, we do DQA…”* (IDI1 subnational level)

This quote illustrates how subnational managers used donor support to fulfill the requirements of the national agenda on quality assurance in the face of unpredictable *basket funds*, the main financial source for implementation. Since donor support was not ubiquitous, this strategy sometimes caused clashes, when the respective donor also supported the introduction of parallel data collection systems.

### 4.2. Theme 2: Shaping HMIS design through decisions

#### 4.2.1. No money, no plan

Lack of financial power influenced the frequency of physical DQA where primary data review and bonding with data collectors took place while providing and receiving feedback in person. Managers explained how funds from the basket fund were late or less than budgeted. Prioritization did therefore not necessarily include HMIS despite the overall agenda.

> *“You know in the health sector if you go to prioritize hiring employees, they will prioritize doctors, nurses, pharmacists. But if you tell them about ICT [Information and Communication Technology], they say “Aaaah! just wait, next financial year”, every time next financial year*.*”* (IDI 9 national level)

This situation made activity implementation difficult. It is also very likely that these experiences informed subnational managers’ different agenda related to data sovereignty, cooperating closely even with donor organizations whose agenda did not match the national-level strive for data sovereignty.

#### 4.2.2. Implementing data systems

National-level managers depicted how they designed digitalization iteratively considering infrastructural, workforce and financial constraints.

> *“There is no standard formula so “try and error” with [digital] systems is done… Everybody is eager to go digital, but how is unclear*.*”* (IDI 16 national level)

Some respondents explained that health data system design was new in the African context making it hard to find adequate examples for roll-out.

Conceptualizing DQA in contrast seemed easier, since WHO had developed guidelines and tools in the past, now included in DHIS-2. These DQA tools were the main lever for data quality for national-level managers. Their data verification strategy relied on i) statistical programmes, ii) personal analytical skills, and iii) triangulation with external (numeric) data sources and within multi-professional teams.

Subnational managers had more opportunities to “know data and how it is collected” due to their physical proximity to facilities, making this their main lever of control. Their i) clinical background, ii) hands-on experience with data collection and iii) knowledge of working conditions and infrastructural challenges in health facilities, helped them triangulating data to develop context-specific implementation strategies, often driven by the lack of resources.

> *“Planning means using those resources that are already there. This is why we must adjust according to reality. If we are supposed to visit ten facilities, we should at least visit three or four [facilities] depending on the amount that has been sent*.*”* (IDI 7 subnational level)

Political leaders had committed to data-driven policymaking, and data-entry itself was mandatory. National managers thus used mainly institutional power for HMIS design and policymaking including enforcement measures for accountability and quality assurance. Working around funding and contextual challenges however, often demanded use of discretionary power.

#### 4.2.3. Enforcing accountability

Accountability processes were included into HMIS design. These were: i) establishing a clear line of reporting, ii) negative consequences for non-compliant individuals, iii) providing timelines and following up, iv) adding signature or phone numbers on data reports and v) establishing data review spaces where appointed individuals reported. Accountability through signature mainly concerned facility managers ensuring that data leaving facilities was acceptable.

> *“I personally think this needs sensitization starting with managers. Because the normal staff, doctor, nurse or anyone may have produced the data*… *You know, the data will not leave the facility without being seen by the in-charge. Now these supervisors themselves should be the ones if their staff has not seen it, then they should see that the data is not ok*.*” (IDI 13 national level)*

The importance of these facility managers as gatekeepers for data quality and use may be underestimated. They often compiled facility data summaries from registers. While formally managing the facility, they still shared experiences around the precarious environment of data collection with HCPs.

Subnational managers’ main lever ensuring accountability from facilities was the monthly data review before digitization of paper-based data. Previously data was verified against monthly summary reports, but now districts had changed to using facility registers. Subnational managers reported spending much time on data validation and given that they too had submission deadlines, it is likely that some of the above measures may have been skipped. In addition, many of the described processes could in theory be negotiable and national managers knew that DQA partly depended on funds. This situation may have weakened the use of institutional forms of power to enforce accountability, hence the high hopes set on digitalization.

### 4.3. Theme 3: Shaping HMIS design through connection

#### 4.3.1. Being self-motivated

Managers, especially at national level, described feeling motivated by knowing they contributed to a greater good.

> *“I am very motivated when I see that data I have looked at and managed is used at the highest level, when they announce something, and this (data) has passed through my hands*.*”* (IDI 10 national level)

It is not self-evident that HCPs felt equally driven and empowered as their managers, to change health data systems for the better as other factors than internal motivation may have influenced their incentive. Managers thought that few HCPs understood the importance of data and statistics for their work, despite using them for annual planning.

> *“I think [data culture] is a variable practice…What I say, where I say that. Much depends on the senior managers there. How do they look at data? How much do they see data as an important variable?”* (IDI 11 national level)

Managers’ own experiences were thus deemed important for HCPs to develop a positive view on data. Many managers shared an often-empathic image of HCPs with regards to their capacity and motivation to collect data and use it. Managers also acknowledged that HCPs had a high workload and could thus lack time for data processing.

> *“Sometimes you can say “maybe I should do a certain task” but if you look at the shortage of people in facilities and the amount of work, you will find them saying “I will do this [task] later and they forget*.*”* (IDI 4 subnational level)

Other managers attributed HCPs’ problems with digital data collection rather to human behavior than lack of technology affinity, i.e. i) a natural resistance to change, and ii) lack of interest of HCPs in data.

> *“It is not that they are backward, it is human behavior, and this is really a big problem. There are places where you install a system, and they don’t use it at all, and you ask yourself why. Because these things start with someone’s interest and willingness to change what they were used to previously*.” (IDI 10 national level)

Overall HCPs were perceived as lacking agency to collect data correctly to be used by others or themselves and seemingly managers had no working hypothesis how to solve this.

#### 4.3.2. Communicating with different actors

Subnational managers used diverse communication channels to form relations and influence implementation, while national managers employed more hierarchical, formalized spaces for communication.

National-level communication consisted mainly of top-down feedback around erroneous data. In contrast subnational managers frequently communicated in various directions within the system. Communication with the national level was done through WhatsApp groups or by mail for more serious issues. These groups, formally installed as digital spaces for data review, played a prominent role in their communication on HMIS matters.

> *“Yes, and some of them [district teams] when they do those [data review] meetings, they invite us and send pictures like:” Guys, today we had a meeting, we did one, two, three*.*”* (IDI 7 subnational level)

This quote emphasizes the relational role these groups played at subnational level, but also how accountability was displayed upwards through this group.

#### 4.3.3. Negotiating accountability

Most managers described accountability as subjected to natural occurrence, generational change or following digitalization rather than something that could be groomed strategically. Managers however, mentioned how they negotiated HCPs’ accountability through i) leadership or role modelling, ii) experiential learning, iii) social pressure, iv) sensitization or formal training and v) financial motivation. Subnational managers described how they proactively used peer or social pressure to negotiate accountability from HCPs.

> *“Yes, you feel shame, even guilt*…*Sometimes [we ask them]” Imagine that thing you did, it was done to you, how would you feel? They feel guilty by themselves, so it is obvious they must change*.*”* (IDI 4 subnational level)

This quote underlines how strongly the lever of “being seen” may be perceived regarding accountability. This may also explain why respondents held such expectations towards digitalization, where people and performance are made visible in the digital space.

### 4.4. Overarching theme: Shaping HMIS governance using stick and carrot

Managers described how they used different forms of power to set the national HMIS agenda and implement it against the background of a highly complex context. Political and institutional power transmitted a clear data quality agenda at all levels. This enabled national managers to install a set of rules related to implementation of DQA, i) the use of WHO software, ii) regular physical DQA at facilities.

Institutional power use was thus successful in an area where aim and means were clearly outlined along the line of command. In the area of data use, the aim was equally well outlined, but not the means to reach that goal. This uncertainty may have provided room for discretionary power use regarding directions to lower levels, e.g. on whether training or experiential learning through use should be applied, but also regarding implementation, e.g. the degree of support that facilities received for data analysis.

Many of the data verification and monitoring processes were delegated to subnational managers but national managers trusted the data they managed and information they passed on to politicians, mainly because they trusted the processes they had institutionalized.

> *“I am confident, not because I am confident that the data is best [quality]. I am confident because this data goes through all the processes until it reaches me*.*”* (IDI 10 national level)

National managers had thus delegated important power to their colleagues at subnational level. Some parts of the national agenda however were seemingly translated differently through discretionary power at subnational level, e.g. on data use or regarding data sovereignty, sometimes leading to i) unintended outcomes, e.g. the creation of donor-driven data silos, or ii) no outcomes, e.g. lack of data use for service improvement at facility level. But discretionary power practices also led to innovative approaches to implementation at subnational level. HCPs, in contrast, were seen as actors driven by human nature and difficult to change, who seemingly needed more control. Innovations at that level may consequently have been overlooked.

## 5. DISCUSSION

Our findings suggest that managers working in HMIS governance in Tanzania have adapted much of their agenda-setting and decision-making on governance and implementation to the complex dynamics of their context, using i) institutional power where guidance was clear and less uncertainty existed, e.g. in the case of DQA, and ii) discretionary power in more unpredictable fields such as data use. In addition, managers’ different positionalities with regards to data and data systems in general may have contributed to the application of different forms of power to interpret and implement policy and agendas. Health care providers were perceived as unpredictable agents for data quality and use, thus accountability was mostly negotiated using discretionary power through connection and positionality and, where possible, institutional power through processes enforcing accountability such as signatures and data verification at different levels.

Availability of international tools such as the WHO DQA tool kit, and contextual challenges, e.g. HCPs’ lack of agency and accountability, shaped national agendas related to HMIS governance and design: There were few uncertainties on how to scale-up DQA and consequently institutional power was sufficient to transport this agenda to the lower levels with a simple set of rules (22). Implementation was mainly hampered by funding issues, so subnational managers had to make discrete choices to make things work, but they seemingly did this within the boundaries set by their superiors and national policies.

Although data use was also part of international discussions around HMIS strengthening (41, 42), there was no consensus yet on the modalities of implementation. The fact that the most prominent data-use-frameworks do not define the link between improved data generation and use (8, 41, 43) may underline this lack of consensus on leveraging factors despite an ample body of literature testing data use interventions (8, 9, 44-46). Our results suggest that uncertainty around purposes and means to improve data use was translated into national agendas and consequently their application. This created opportunities for discretionary power practices at all levels.

We found that some policy aspects got lost in discretionary translation down the line of command, e.g. regarding data sovereignty, similar to Lehman et al. and others, describing how discretionary power used by *street-level bureaucrats* transformed policy at subnational and facility level (31, 47, 48). In contrast to Lehman et al., Carlitz et al. stress how health care managers led Tanzania’s COVID-19 strategy through discretionary innovation against the background of lacking policies (30). We also concur with this interpretation for the highly adaptive complex HMIS in Tanzania. We identified several innovative approaches to interpret national agendas or policies, e.g. data verification measures at subnational level and use of remote platforms for relationship-building to increase contact time despite financial unpredictability.

Our research findings further illustrate the influence of health system bottlenecks such as i) financial constraints, ii) human resource problems and iii) lack of supervision and feedback, on the use of different power forms in HMIS governance and implementation (20). This is in line with previous findings from the evaluation of a digital support tool for HMIS data collection at facilities in southern Tanzania and other research on data quality within the HMIS (9, 13, 23). The resulting unpredictability, also documented for clinical care in the Tanzanian setting (49, 50), prompted especially subnational managers to apply discretionary power so they could still deliver, e.g. choosing easy-to-reach facilities for DQA, instead of others where physical visits may have needed more resources. These findings underline the interdependency of the overall health and HMIS systems as described for complex adaptive systems (25, 39).

Our findings indicate that HMIS governance in Tanzania and system adaptations are shaped by relational interaction of different actor groups as described for complex adaptive systems (22, 25, 39). Consequently, actors’ agency, i.e. their capacity to take action, seemed an important factor to move implementation into the right direction (22). Our results further suggest that current HMIS design rather reflects national managers’ motivators (22) instead of considering all actors within the system: Managers’ motivators were mainly internal since they were working at a health system level where the grand picture of well performing data systems may be more easily perceivable. Studies from similar settings suggested that HCP, in contrast, feel they are collecting data for someone else, partly because they do not use it themselves (14, 15, 51). Estifanos et al. report that rewards to facilities are often driven by data-based performance only in Ethiopia, which increases incentives for data manipulation in a working environment were tools to create this performance are often lacking (15). We have previously reported that HCPs in maternity wards in southern Tanzania used data to safeguard social relationships with i) immediate supervisors, ii) subnational and national managers and iii) the community, which are otherwise difficult to maintain in the given environment (12). We also reported that the value HCPs’ assign to data collection in different formats may differ from their supervisors’ (23), partly because the former need a different type of information than collected via DHIS-2 (12). Our current findings indicate that health care managers perceived HCPs as unreliable in terms of their aptitude to produce quality data and use them. Consequently, innovations at this level may have gone unnoted. Our previous research evaluating the introduction of a novel hybrid-digital routine health data collection tool suggests nonetheless, that HCPs develop innovative approaches to embed digital tools and improve data collection despite health system challenges (23). We therefore propose that complexity and interdependency of routine health data collection and health care itself should be acknowledged as a first step when thinking about data collectors’ sense of agency and about what motivates them.

We found that managers consider agency and accountability a human behavior that is difficult to predict or enforce. Discretionary power practices at subnational level including the negotiation of accountability may be a more suitable tool than enforcing it in the given context (22, 52), but we argue that this discretion could be better directed by institutionalization of a set of organizational values related to data collection and processing which could guide i) agenda setting, ii) communication between actors and iii) implementation. Also here, facility in-charges may play a crucial role to ensure that HCPs’ motivators are sufficiently considered in this process, but also to monitor outcomes and recommend adaptations over time.

### 5.1. Methodological consideration and limitations

The research team has extensive experience in stakeholder management from different angles and thus a high understanding of the context in which our participants operated. Still, social desirability bias could have been at play among managers related to i) superiors, ii) the public and iii) politicians. We tried to limit this possibility by using external gatekeepers from international organizations to identify potential participants. Extensive peer check was used during data analysis to identify areas in the data set where social desirability may have guided the elaborations.

## 6. CONCLUSIONS

Health care policymakers at all health system levels experienced challenges in health management information system governance related to unpredictability of funding and unreliable performance of health care providers, a core actor group in the system. They resorted to different strategies depending on perceived uncertainty levels to resolve these.

Policymakers and senior managers need to acknowledge the interconnectedness of HMIS and the overall health care system to achieve an alignment between policy and implementation. This should also include reflecting on prerequisites for all actor groups’ ability to perform well, like the availability of funds, (clinical) working tools and a set of minimum specifications to guide policy implementation.

We further suggest the importance of identifying a set of social norms and values around data collection and processing that resonate with all actor groups and may support governance of this complex system. Subnational and facility managers may be in the best position to observe these values’ influence on implementation but are currently not capacitated to do so.

## Data Availability

All relevant data are within the manuscript. Qualitative primary data such as transcripts reflect views of managers from district, regional and national level. Since the HMIS community in Tanzania is very small, and the number of subnational managers is low, individual participants are thus easily identifiable. Making the full data set publicly available could potentially breach the privacy that participants were promised upon request for participation. Also, our ethics approvals from Muhimbili University of Health and Allied Sciences and National Institute for Medical Research′s Ethics Committee were granted based on the anonymity of the individuals consenting to participate. Due to these conditions, the authors are unable to avail the full transcripts. Excerpts of specific segments of the text will be reviewed for any potentially identifying details and made available to fellow researchers or reviewers who complete a data sharing agreement and abide by strict confidentiality protocols. In line with the information given to the participants and restrictions set by the ethics committees above, access to the full transcripts is only available to the involved researchers. Data requests may be sent to the corresponding author, RU, via.

